# CT-based Machine Learning for Donor Lung Screening Prior to Transplantation

**DOI:** 10.1101/2023.03.28.23287705

**Authors:** Sundaresh Ram, Stijn E Verleden, Madhav Kumar, Alexander J. Bell, Ravi Pal, Sofie Ordies, Arno Vanstapel, Adriana Dubbeldam, Robin Vos, Stefanie Galban, Laurens J. Ceulemans, Anna E. Frick, Dirk E. Van Raemdonck, Johny Verschakelen, Bart M. Vanaudenaerde, Geert M. Verleden, Vibha N Lama, Arne P. Neyrinck, Craig J. Galban

**Author notes:** Corresponding author Sundaresh Ram, PhD, Department of Radiology, University of Michigan, Biomedical Science Research Building, Room A205, 109 Zina Pitcher Place, Ann Arbor, MI 48109-2200. (co-first author). (co-last author).

## Abstract

**Background:** Assessment and selection of donor lungs remains largely subjective and experience based. Criteria to accept or decline lungs are poorly standardized and are not compliant with the current donor pool. Using ex vivo CT images, we investigated the use of a CT-based machine learning algorithm for screening donor lungs prior to transplantation.

**Methods:** Clinical measures and ex-situ CT scans were collected from 100 cases as part of a prospective clinical trial. Following procurement, donor lungs were inflated, placed on ice according to routine clinical practice, and imaged using a clinical CT scanner prior to transplantation while stored in the icebox. We trained and tested a supervised machine learning method called *dictionary learning*, which uses CT scans and learns specific image patterns and features pertaining to each class for a classification task. The results were evaluated with donor and recipient clinical measures.

**Results:** Of the 100 lung pairs donated, 70 were considered acceptable for transplantation (based on standard clinical assessment) prior to CT screening and were consequently implanted. The remaining 30 pairs were screened but not transplanted. Our machine learning algorithm was able to detect pulmonary abnormalities on the CT scans. Among the patients who received donor lungs, our algorithm identified recipients who had extended stays in the ICU and were at 19 times higher risk of developing CLAD within 2 years post-transplant.

**Conclusions:** We have created a strategy to ex vivo screen donor lungs using a CT-based machine learning algorithm. As the use of suboptimal donor lungs rises, it is important to have in place objective techniques that will assist physicians in accurately screening donor lungs to identify recipients most at risk of post-transplant complications.

## Introduction

Lung transplantation continues to be the only treatment option for many patients with end-stage lung disease. Its success remains limited by the discrepancy between the number of patients on waiting lists and the availability of donor organs, resulting in significant waitlist mortality (approximately 10% of lung transplant candidates within the Eurotransplant network).^1^ Therefore, options to increase the donor pool, based on well-implemented extended donor criteria, are being explored. Nevertheless, the lung recovery rate of a multiorgan donor remains limited to 20%-30% in most centers.^1,2^ In order to overcome this shortage, it is crucial that we critically evaluate our current practices in assessing organs prior to transplantation. At this moment, there are only moderate evidence-based criteria for donor lung assessment, based on a combination of donor history, clinical parameters (e.g., gas exchange), chest X-ray, bronchoscopy findings, and ultimately, in situ visual inspection by the transplant surgeon.^3-8^ Donor lung acceptance remains largely subjective and dependent on macroscopic appearance and expertise of the surgeon.^9-11^

In 2017 we evaluated the use of high-resolution X-ray computed tomography (CT) to assess donor lungs, potentially increasing the pool of high-quality lungs for transplantation.^12^ This study evaluated the use of CT to radiographically assess the presence of lung abnormalities. We found that many lungs declined for transplantation showed no obvious signs of disease or injury based on CT screening, which suggests they were adequate for transplantation. In a subsequent study, we critically assessed reasons for not using donor organs for transplantation by in-depth CT and histopathologic assessment and showed significant discrepancy between clinical indication for not using the organ for transplantation and quality of the lungs as shown on CT. This clearly illustrates the need for another tool to critically assess donor organ quality before transplantation.^13^ Other groups have also investigated the potential utility of CT assessment of donor organs. Gauthier et al. leveraged in vivo chest CT scanning by demonstrating its value for determining the presence of structural lung injury such as emphysema as a tool for screening a large group of potential donors.^14^ Bozovic et al. also compared information derived from standard lung X-ray screening to chest CT imaging and found that a targeted imaging review of abnormalities affecting the decision to use donor lungs may be useful in the preoperative stage.^15^ In a separate study, Sage et al., using real time CT imaging, was able to monitor improvements in lung parenchyma during ex vivo lung perfusion, a tool that assesses and potentially reconditions donor organs prior to transplantation.^16^ While this experimental data demonstrates an added value of chest CT scanning in donor assessment and selection, adopting CT for screening may be hindered by availability of trained thoracic radiologists and increased wait times during assessment of the donor lungs in a process that is critically time-dependent.

Machine learning (ML) is a branch of artificial intelligence where a computer algorithm learns from examples to generate reproducible predictions and classifications of previously unseen data. Once trained, this computational technique can be automated to analyze large amounts of data in a relatively short period of time. ML continues to be extensively investigated for tissue/organ segmentation, prediction, and classification in a wide array of medical imaging applications including transplant medicine.^17,18^ Specifically, supervised ML in the context of CT lung imaging has been used to detect and quantify airway patterns in pediatric patients with cystic fibrosis,^19^ as well as classify COPD patients based on the Fleischner Score.^20^ These ML models, referred to as “deep learning,” require large data sets for training and testing. When training data is limited and/or noisy, as is often the case in medical imaging, these methods tend to show a performance degradation.^21^ In contrast, ML models known as “dictionary learning” are based on the concept of sparse representation-based classification. The benefit of this ML model is that it assumes each region of the lung in the CT scan, i.e., patch, can be accurately represented as a linear combination of very few elements of the dictionary.^22^ This allows dictionary learning-based models to perform with high accuracy from relatively small datasets.

Incorporating the precision of an ML model into donor lung assessment may have significant clinical impact by preventing the rejection of viable lungs. Potentially improving the accuracy of decisions made by clinicians, moreover, may result in life-saving consequences for patients. We hypothesize that a supervised “dictionary-learning” ML model, applied to ex-situ CT scans of freshly procured human donor lungs, can provide meaningful results that aid in donor lung screening. We developed and investigated an ML algorithm for classifying donor lungs for transplantation that learns to associate unique CT image features that are specific to “accepted” or “declined” lungs as described by thoracic surgeons, without any prior knowledge of the donor or recipient.

## Materials and Methods

### Ethics statement

This study was carried out in 100 subjects enrolled as part of a single-center prospective trial from 2016 to 2018 and was approved by the Institutional Review Board of KU Leuven and University Hospital Leuven (S59648 / B322201630218). It adheres to the principles of the World Medical Association Statement on Organ and Tissue Donation, the Declaration of Helsinki, and the Declaration of Istanbul. Study participation required legal consent to explant declined lungs.

### Design

All potential donor lungs during this period were reviewed on chart by our experienced transplant team for suitability following our routine clinical practice. An initial assessment, based on donor age, clinical history, partial arterial oxygen pressure at 100% fraction of inspired oxygen (FiO2) and 5 cm H2O positive end expiratory pressure, chest X-ray scans, and logistic availability, determined whether a procurement team would be sent to the donor hospital. A donor was considered only when legal criteria of brain death, donation after circulatory death (DCD) III or euthanasia (DCD V) were met, as required by Belgian law. Existing allocation rules were followed.

### Ex-situ Lung Preparation and CT Scanning

In this study, an ex-situ CT scan was taken of every pair of donor lungs after standard procurement. First, the final decision for suitability for transplantation was made by 6 experienced senior thoracic surgeons after in-situ inspection at the donor hospital according to routine clinical practice. Lungs were then flushed (4°C) with cold Perfadex® (XVIVO Perfusion, Gothenburg, Sweden) and inflated with 50% FiO2 at 25 cm H2O. Lungs were packed in cold Perfadex® and stored on ice in a transportation box. Upon arrival at the transplant center, every pair of lungs was CT scanned (Siemens Somatom scanner, Erlangen, Germany) at 120 kV and 110 mAs within the transportation box (static cold storage). The transplant team was blinded from CT information and therefore, any abnormal finding on CT did not influence the decision to proceed with lung transplantation. Inclusion criteria for the study were first single-organ transplantation, successful procurement and legal consent to explant declined organs. Illustration of the workflow and representative CT slice orientations for a donor lung are provided in **Figure 1**.

**Figure 1:**
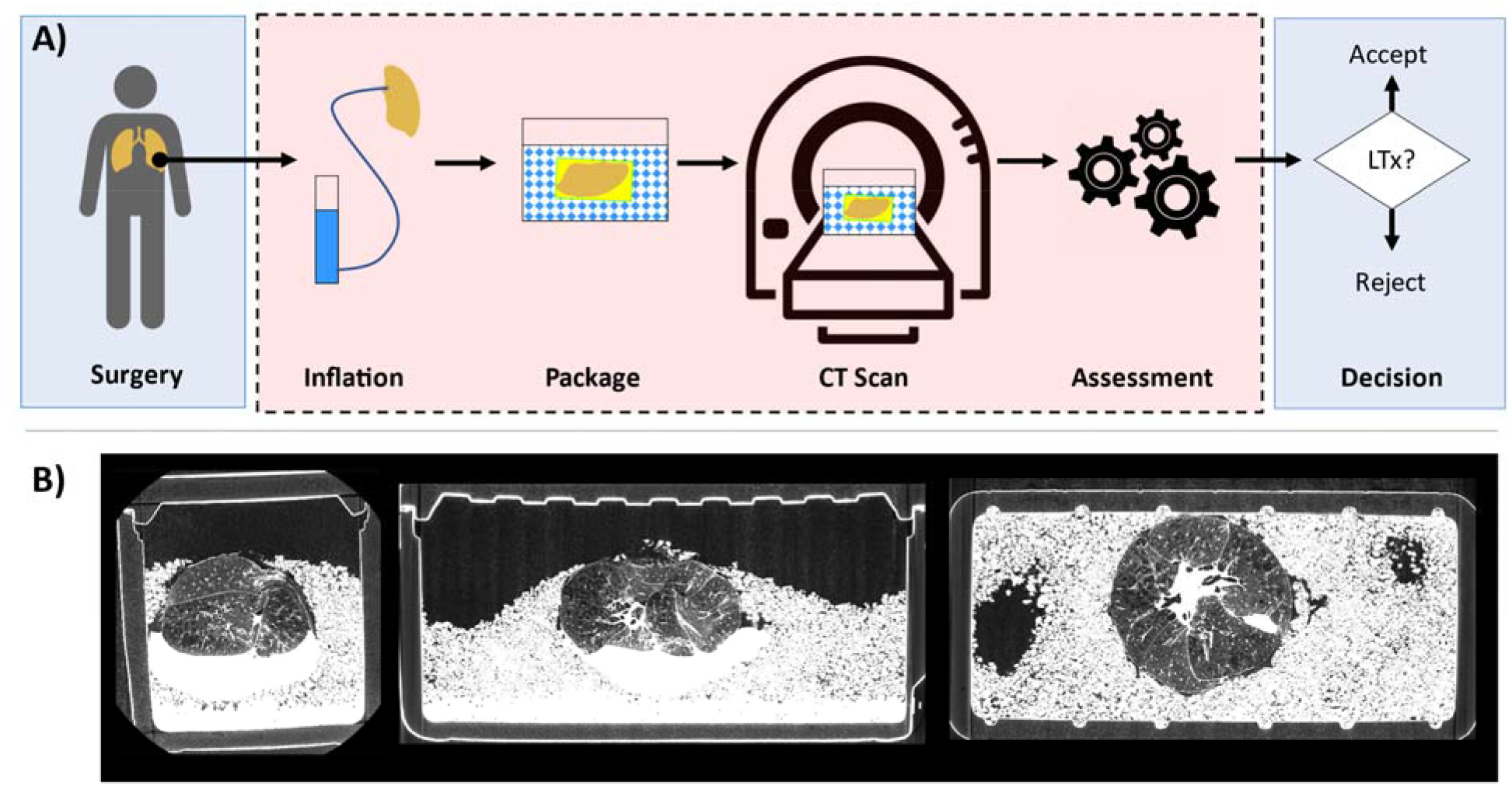
Illustration of donor lung screening with computed tomography workflow. (A) Provided is an illustration of the inclusion of CT in routine donor lung screening process. Blue boxes represent standard-of-care, and red box represents CT-ML procedures. The approximate time for donor lung preparation and CT imaging is 5-10 minutes. (B) Corresponding axial, sagittal and coronal views of a CT scan from a declined donor lung (**Figure 3C-D** and Case 4 in **Table 3**).

### Clinical variables

In all recipients who eventually received the CT-scanned grafts, primary graft dysfunction (PGD) was defined according to the latest ISHLT guidelines.^23^ Clinically relevant parameters were collected from both donors and recipients including age, sex, height, weight, pO2, ventilation time, pulmonary function measurements, PGD, hospital stay, ICU stay and chronic lung allograft dysfunction (CLAD)-free over 2 years. Information on one recipient was not available. The donor lung for this recipient was randomly selected as a test case for evaluation of our ML algorithm.

### Machine Learning Analysis

Using a bespoke automated segmentation algorithm, lungs were segmented to remove the influence of ambient air, ice, and the box on the ML model. Our ML model is a dictionary learning algorithm that classifies CT features from lung tissue as “normal” or “abnormal.” For training of our ML model, “ground truth” was set to the final decision by 6 experienced senior thoracic surgeons as part of routine clinical practice. Training was performed on a randomly selected subset of 14 cases, split evenly between accepted (N=7) and declined (N=7) for transplantation. The remaining 66 cases were used for testing. This subset consisted of 52 accepted and 14 declined donor lungs. In brief, our ML model is designed to associate unique CT features that are specific to “accepted” and “declined” lungs. This is achieved by randomly selecting subsets of CT data (i.e., patches) and comparing the underlying patch features with the compiled class dictionaries of features, which were determined during training. It is important to note that no prior knowledge about the donor, recipient, and lung tissue features, such as emphysema, honeycombing, ground glass opacities or consolidation, were provided for the algorithm to delineate “normal” from “abnormal” lung tissue. Details on model design and methods for training and testing are provided in the Supplement (**Supplemental Figure 1, Methods, and Results**). All processing and analyses were performed using in-house algorithms developed in MATLAB version 2020a (MathWorks, Natick, MA).

### Statistics

Continuous and categorical variables were expressed as mean ± standard deviation and total number and percentage, respectively. For transplanted lungs identified by ML as “Declined” (N=13) and “Accepted” (N=39), differences in continuous and ordinal variables were analyzed for statistical significance using a Mann-Whitney U test. Categorical variables were analyzed using Pearson chi-square test. Separate analyses were performed for the highest PGD score. PGD score was used to stratify cases by values <3, classified as 1, and equal to 3, classified as 0. The risk assessment of a donor lung transplant identified by ML as “declined,” resulting in a PGD score of 3, was determined by calculating the odds ratio. Same risk analysis was performed for CLAD-free at 2 years. The extent of ICU stays for donor lung recipients was evaluated using a Kaplan-Meier plot and a long-rank test. Statistical work was undertaken using MATLAB R2019a, and IBM SPSS Statistics v27 (SPSS Software Products). In all tests significance was defined by p < 0.05. For clinically declined lungs (N=14), reason for decline was evaluated in lungs identified by ML as “Declined” (N=9) and “Accepted” (N=5).

## Results

### Subject Characteristics

Of the 100 donors identified between 2016 and 2018, we were able to generate adequate lung segmentation from 80 cases, of which 59 were accepted and 21 were declined for transplantation. Provided in **Table 1** are donor characteristics and relevant metrics for transplantation. Donor lungs used for transplantation originated more often from males.

**Table 1:**
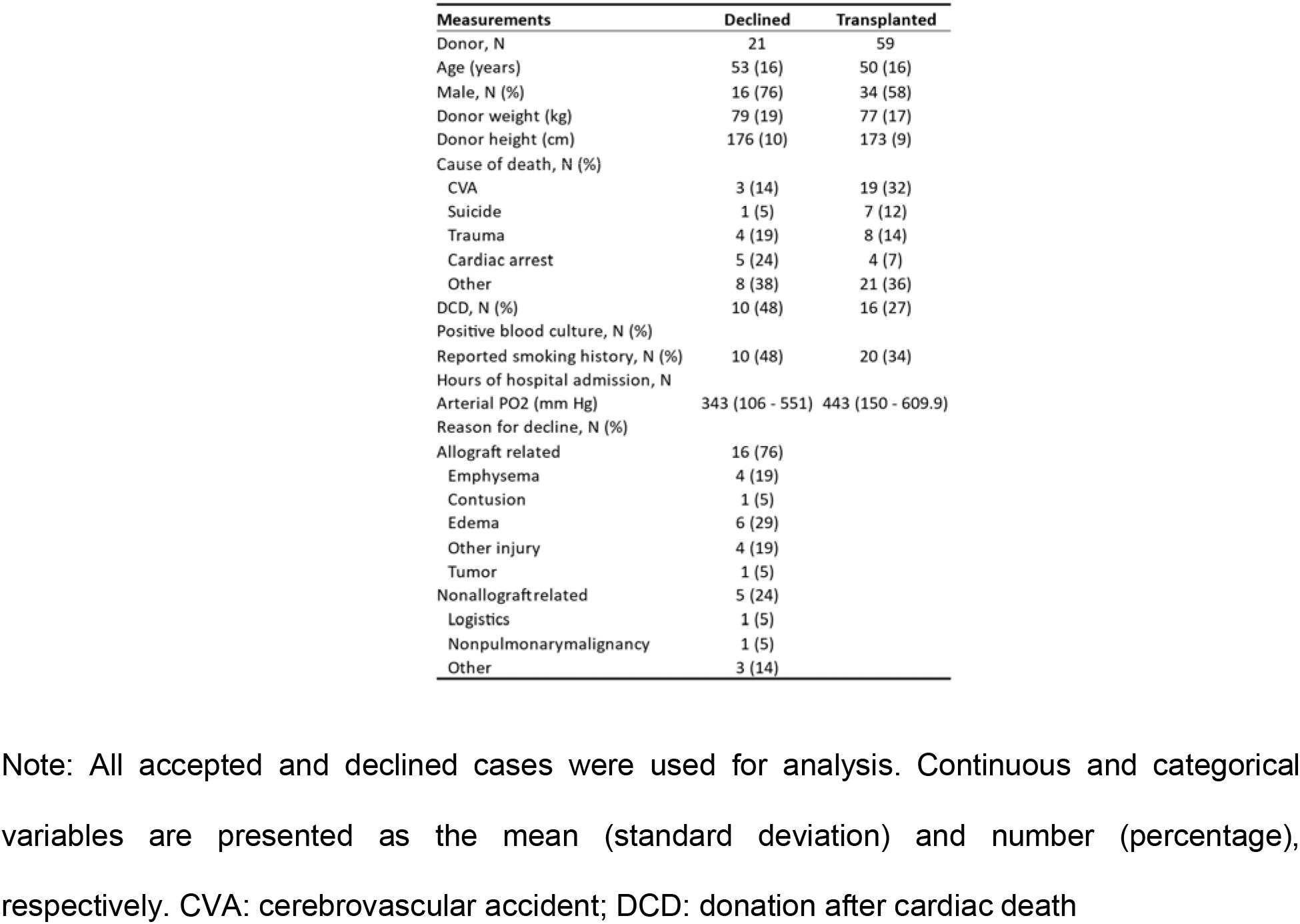
Donor Characteristics

| Measurements | Declined | Transplanted |
| --- | --- | --- |
| Donor, N | 21 | 59 |
| Age (years) | 53 (16) | 50 (16) |
| Male, N (%) | 16 (76) | 34 (58) |
| Donor weight (kg) | 79 (19) | 77 (17) |
| Donor height (cm) | 176 (10) | 173 (9) |
| Cause of death, N (%) |  |  |
| CVA | 3 (14) | 19 (32) |
| Suicide | 1 (5) | 7 (12) |
| Trauma | 4 (19) | 8 (14) |
| Cardiac arrest | 5 (24) | 4 (7) |
| Other | 8 (38) | 21 (36) |
| DCD, N (%) | 10 (48) | 16 (27) |
| Positive blood culture, N (%) |  |  |
| Reported smoking history, N (%) | 10 (48) | 20 (34) |
| Hours of hospital admission, N |  |  |
| Arterial PO2 (mm Hg) | 343 (106 - 551) | 443 (150 - 609.9) |
| Reason for decline, N (%) |  |  |
| Allograft related | 16 (76) |  |
| Emphysema | 4 (19) |  |
| Contusion | 1 (5) |  |
| Edema | 6 (29) |  |
| Other injury | 4 (19) |  |
| Tumor | 1 (5) |  |
| Nonallograftrelated | 5 (24) |  |
| Logistics | 1 (5) |  |
| Nonpulmonary malignancy | 1 (5) |  |
| Other | 3 (14) |  |

Note: All accepted and declined cases were used for analysis. Continuous and categorical variables are presented as the mean (standard deviation) and number (percentage), respectively. CVA: cerebrovascular accident; DCD: donation after cardiac death

### Representative Cases

Presented in **Figure 2** are representative CT slices with the corresponding patch probabilities overlay for two cases: one accepted (**Figure 2** top row) and one declined (**Figure 2** bottom row) for transplantation. The patch probabilities represent the likelihood that the lung tissue within the patch is “normal” (red with probability of 1) or “abnormal” (blue with probability of 0). The donor lung used for transplantation, obtained from a male non-smoker (45-50 years old), was found to consist primarily of patches with high probabilities of normal lung tissue (**Figure 2B**). In contrast, the donor lung declined for transplantation, obtained from a male with over 20 pack years smoking history (65-70 years old), was found to have extensive emphysema (**Figure 2C**) associated with low probabilities of normal lung tissue (**Figure 2D**).

**Figure 2:**
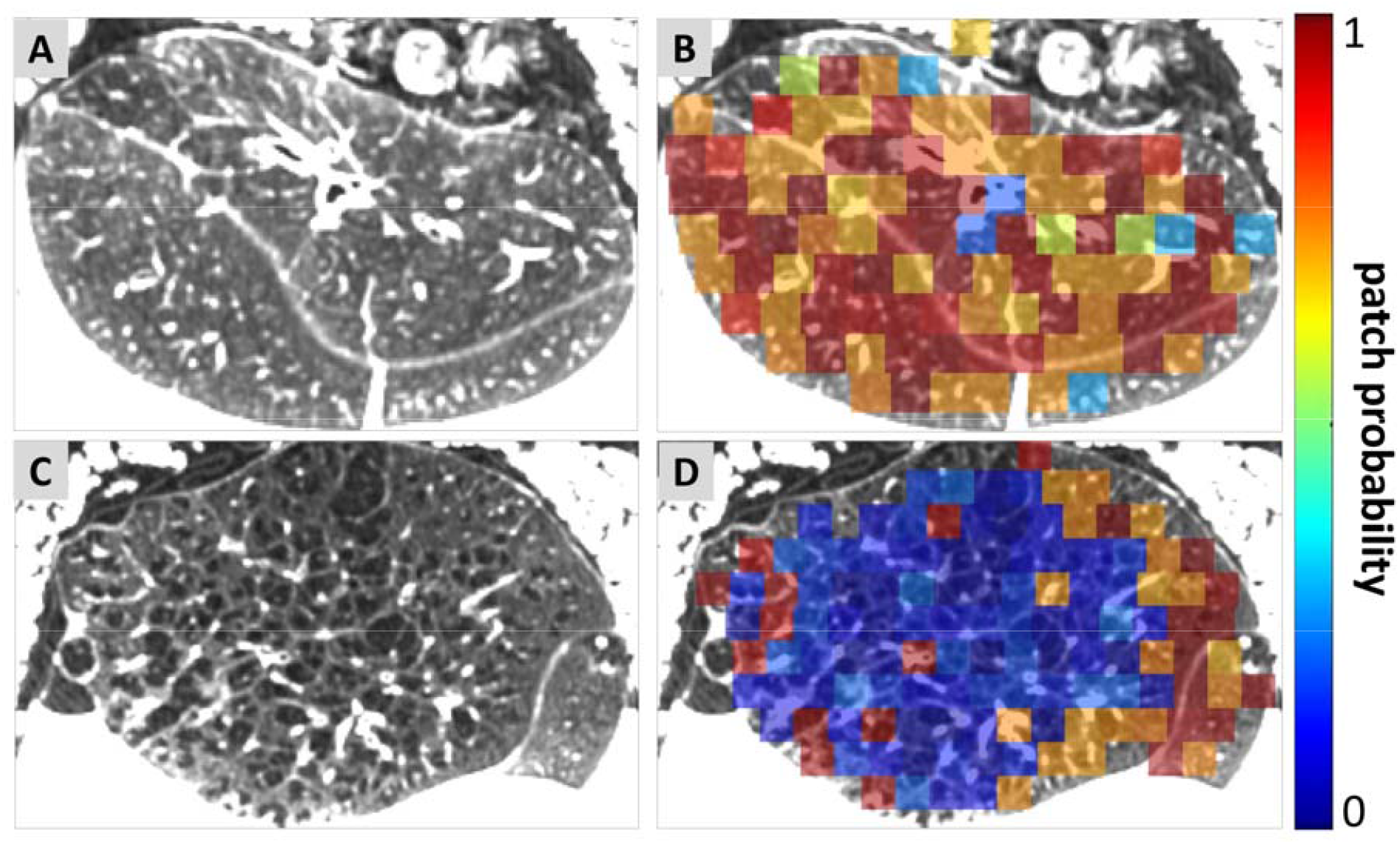
Representative CT scans with corresponding ML patch probability maps for (A and B) accepted and (C and D) declined donor lungs. The patch probabilities represent the likelihood that the lung tissue within the patch is “good” (red with probability of 1) or “bad” (blue with probability of 0). The accepted donor lung was obtained from a male, non-smoker, 45-50 years of age. The declined donor lung was obtained from a male, over 20 pack years, 65-70 years of age, found to have extensive emphysema.

### Accepted for Transplantation

Although our model was trained on the final decision for transplantation, we observed a high number of false positives and negatives (**Supplemental Figure 3)**. Of the 52 donor lungs found to be acceptable for transplantation, around 20% were predicted to be unacceptable (i.e., declined by the model; hereafter “ML Declined”). As shown in **Table 2**, ML Declined donor lungs had feature probabilities significantly lower (0.205 +/- 0.042) than those accepted by the model (“ML Accepted”) for transplant (0.637+/-0.134, p <0.0001). Stratifying the donors based on our model’s predictions, we found no significant differences in donor or recipient characteristics (**Table 2**). Nevertheless, post-transplant outcomes of recipients were found to differ between model predicted groups of transplanted lungs. Hospital and ICU stay post-transplant were both found to be significant (p = 0.039 and 0.0004, respectively), whereas days until extubation and serial FEV1 and FVC (**Supplemental Table 1**) were not. Kaplan Meier plot (**Figure 3**) showed that recipients that received an ML Accepted donor lung had a median ICU stay of 9 days, compared to 14 days for ML Declined donor lungs (transplanted). Dichotomizing recipients based on PGD = 3 and PGD < 3 generated a p value of 0.034 and an odds ratio of 5.23 (95% confidence intervals of 1.02 to 26.73). This implies that a recipient with an ML Declined donor lung is 5.23 times more likely to have a PGD score of 3 than if that recipient had an ML Accepted lung. In addition, recipients that received a ML Declined donor lung were 19.13 (95% confidence intervals of 3.98 to 91.80) times more likely to develop CLAD within two years than their ML Accepted counterparts.

**Table 2:**
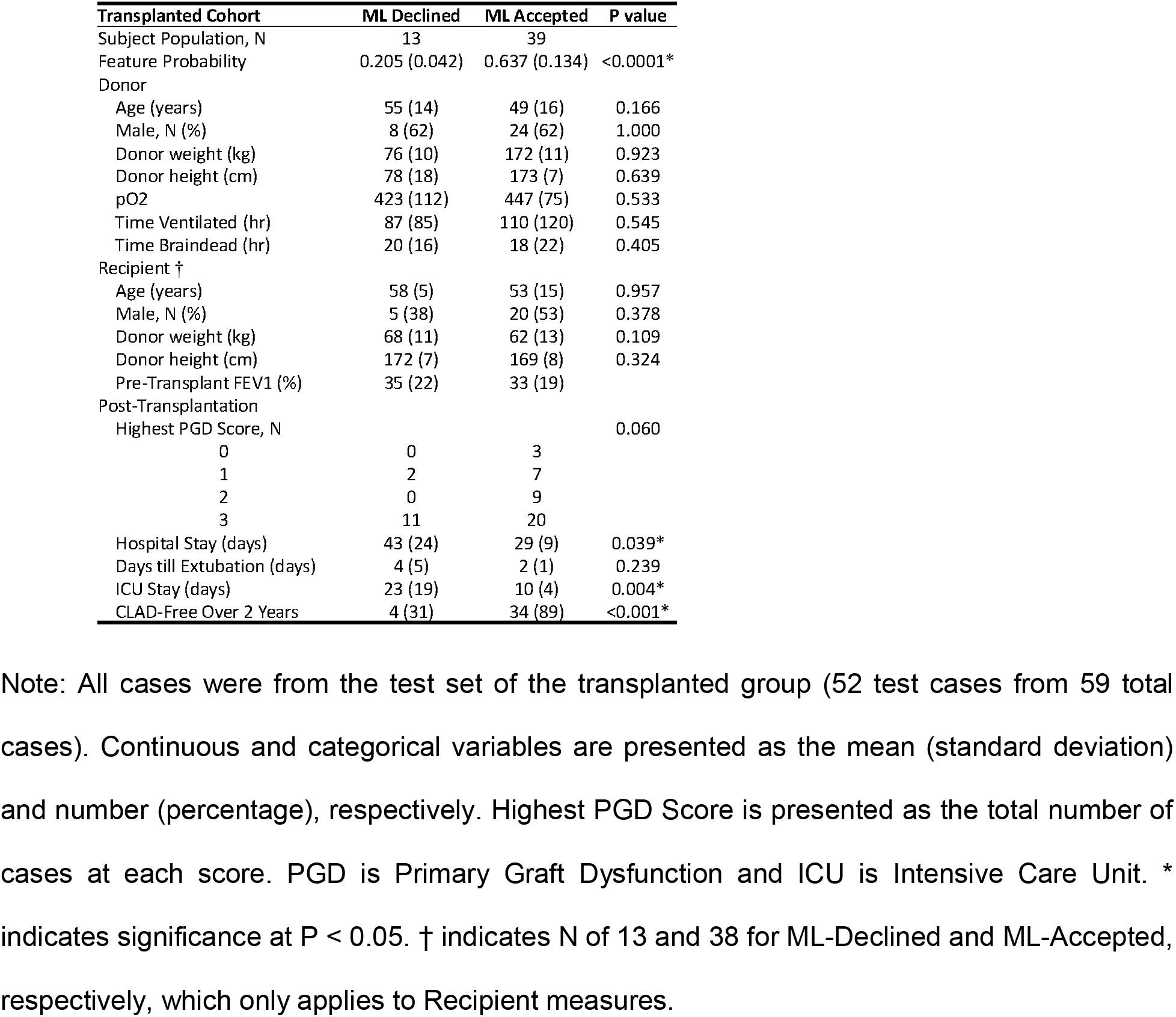
Donor and Recipient Characteristics and Post-Transplant Metrics

**Figure 3:**
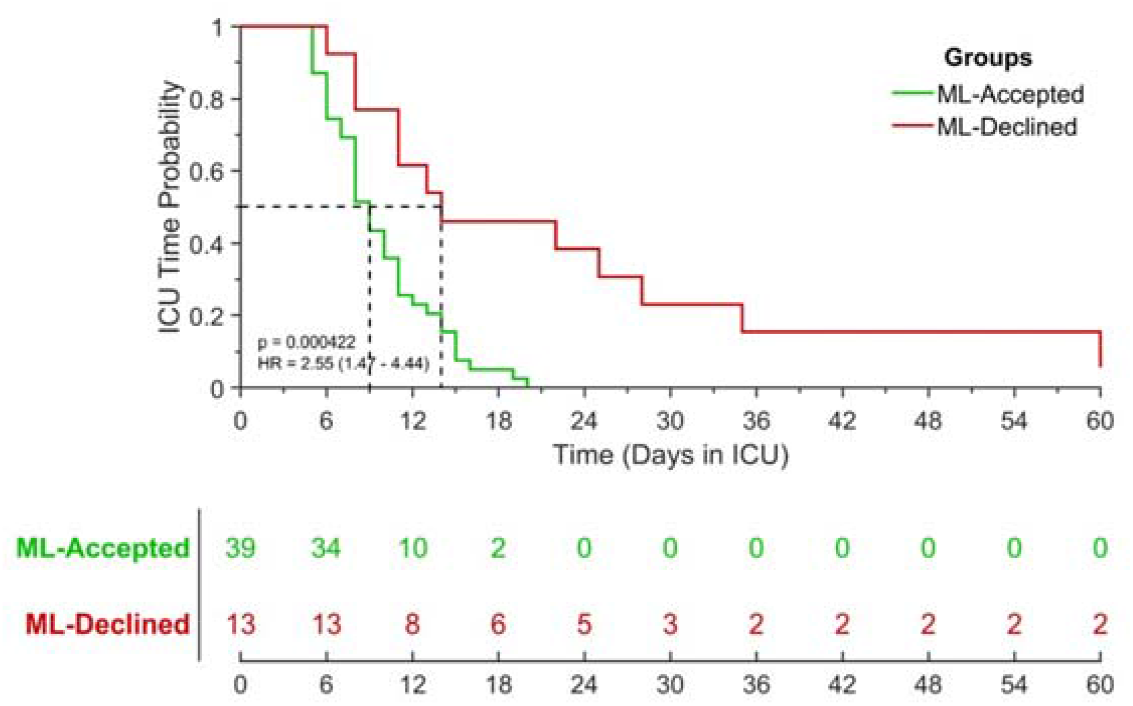
Kaplan-Meier plot showing potential of CT-ML strategy to predict ICU stay in lung transplant recipients (N=52). Green line and red dashed line represent agreement and disagreement, respectively, ML model to clinical decision. Lines correspond to color in confusion matrix (**Supplement Figure 3**). Statistical significance was determined using a log-rank test.

### Declined for Transplantation

Of the 14 donor lungs not transplanted, our model demonstrated an agreement of 64% (**Supplement Figure 3**). Feature probabilities between model-identified groups were found to be significantly different (p=0.0005; agreement N=9; 0.205+/-0.027 and disagreement N=5; 0.340+/-0.04). Eight of the nine cases were found to have pulmonary complications ranging from emphysema to pneumonia (**Table 3**). Only Case 7 was declined due to non-pulmonary complications (lymphoma in the liver) and was found to have low probabilities. Cases 10 – 14 in **Table 3** were identified by our model as acceptable for transplantation. Three of the five cases were rejected due to absence of a matching recipient, one case due to pulmonary contusion (70-75 years old), and one case due to pulmonary edema.

**Table 3:** Reason for Decline and feature probability for declined lungs

| Cases | ML Classification | Feature Probability | Reason for Decline |
| --- | --- | --- | --- |
| 1 | 0 | 0.211 | Tumor |
| 2 | 0 | 0.226 | Microtrombi during backflush |
| 3 | 0 | 0.22 | Edema and bronchopneumonia |
| 4 | 0 | 0.205 | Emphysema |
| 5 | 0 | 0.227 | Pneumonia |
| 6 | 0 | 0.185 | Emphysema |
| 7 | 0 | 0.227 | Lymphoma in Liver |
| 8 | 0 | 0.144 | Emphysema and Edema |
| 9 | 0 | 0.201 | Emphysema |
| 10 | 1 | 0.296 | Edema |
| 11 | 1 | 0.352 | No match |
| 12 | 1 | 0.333 | No match |
| 13 | 1 | 0.401 | Contusion and Age |
| 14 | 1 | 0.319 | No match |

Note: ML classification, feature probability and reason for decline provided for each of the test set cases from the declined group (14 test cases from 21 total cases).

## Discussion

Lung transplantation is presently the only viable cure for end-stage lung diseases such as COPD (Chronic Obstructive Pulmonary Disease) and IPF (Idiopathic Pulmonary Fibrosis). In this proof-of-concept study, we demonstrated a strategy to screen donor lungs ex-situ using computed tomography and machine learning. By leveraging the high resolution and air-tissue contrast of CT and enhanced feature-based detection of a machine learning algorithm, we demonstrated the benefits of this unique strategy for lung screening. In our single center study, we found that our method predicted ICU stay and the odds of a PGD score of 3 in transplant recipients. Our results suggest that this CT-ML strategy, which on average takes only 5 minutes, may serve as a complementary step in the screening process of donor lungs for transplantation.

It is important to note that while CT is not the only tool that can assist with transplantation decisions, it has potential as an accessible, valuable method for selecting viable donor lungs. Donor history, blood gases of the pulmonary veins and in situ inspection remain critical factors in clinical decision making; however, in cases where there is uncertainty about the quality of a donor lung, CT scans may reveal insights that facilitate this process.^24,25^ To the best of our knowledge, this is the first study to evaluate the use of CT in conjunction with machine learning to assess donor lungs used for transplantation. This provided a unique opportunity to test the potential of our approach for predicting post-transplant outcomes. In our previous work, we obtained CT scans from declined donor lungs and found that CT examination of these specimens by a trained thoracic radiologist provided detailed information of interstitial changes otherwise obscured during routine donor lung assessment.^12,13^ However, manual screening of CT scans is hampered by interobserver variability, as well as delays due to accessibility to radiologists. Importantly, time constraints must be minimized to effectively incorporate our strategy of applying CT scanning to donor lung screening. For the present study, we therefore developed a fully automated process to screen CT scans of donor lungs.

An important attribute of our ML screening method is its ability to focus exclusively on the features presented in CT scans without requiring additional information such as donor or recipient characteristics or clinical data. Due to the novelty of our method, i.e., using clinical CT scans to screen donor lungs, our data came only from this single center study. While our dictionary learning model was trained only on 14 cases (7 accepted and 7 declined), it still provided associations with clinically meaningful measures. In fact, our model predicted ICU stay in lung transplant recipients (**Figure 3**). Further, we observed significant differences in hospital stay between transplant recipients with donor lungs classified as “accepted” and “declined” (p = 0.039; **Table 2**). PGD scores are used in the early post-lung transplant period (immediately post-transplant to 72 hours post-transplant) to predict early outcomes. Through our strategy of screening donor lungs using CT and ML, we not only demonstrated that recipients who received a “ML Declined” donor lung, as classified by our approach, were 5.25 times more likely to generate a PGD score of 3 but were 19.12 times more likely to develop CLAD in 2 years. It is important to reiterate that no prior knowledge of the donor or recipient, other than the ex-situ CT scan, was used to train our ML model. It is also important to note that the training set, whether accepted or declined, consists primarily of healthy lung tissue. To account for this bias, we developed our algorithm to detect and remove redundancies between dictionaries, such that patches in class 1 comprise of normal lung and class 2 abnormal lung. We identified one case in our training set declined due to logistics, though it was a healthy lung. However, this would not affect our ML algorithm as it would automatically associate normal patches with class 1 irrespective of the case delineation.

### Limitations

There are limitations to the study worth discussing. This study was performed as part of a single center trial. Consequently, CT scans were procured from a relatively small cohort of donor lungs, affecting our statistical power and the amount of data used to train and test our machine learning model. Nevertheless, we were able to overcome this limitation using a “dictionary learning” algorithm based on the concept of sparse representation-based classification. Even with a limited number of cases—N=52 donor lungs accepted and N=14 declined—we were able to demonstrate clinically meaningful results, such as ICU stay in lung transplant recipients (**Figure 3**) and CLAD-free over 2 years. Although our model classifies individual image patches using discrete feature libraries, final classification is performed using all patches and a feature threshold of 0.272 (determined using the ROC plot in **Supplemental Results**). Presented in **Figure 4** is a clinically declined lung identified by our algorithm as acceptable for transplantation (Case 10 in **Table 3**). This donor lung was declined due to edema, clearly seen in the right portion of the image. Evaluation of the patch feature probabilities show that this region of the lung contained abnormalities, but overall, the lung cleared the final classification step with a value of 0.296. In this instance, a trained thoracic radiologist may conclude that the donor lung is acceptable for transplantation. Like all models, there will always be false positives and negatives. Ultimately, our strategy is not meant to replace the current system but to provide additional support to clinicians during the donor screening process that will help them improve patient care and outcome. Inclusion of a map in a final report, like those presented in **Figures 2** and **4**, would assist clinicians in the decision-making process. For this screening strategy to gain acceptance in routine clinical care, we will propose a multi-center prospective trial to evaluate the effect of CT scanner type on ML model performance, which will provide data for improving the lung segmentation algorithm to maintain a fully automated process. Ultimately, we aim to incorporate CT derived information with clinical data from the donor and recipient to assess the overall transplant risk of this donor-recipient combination.

**Figure 4:**
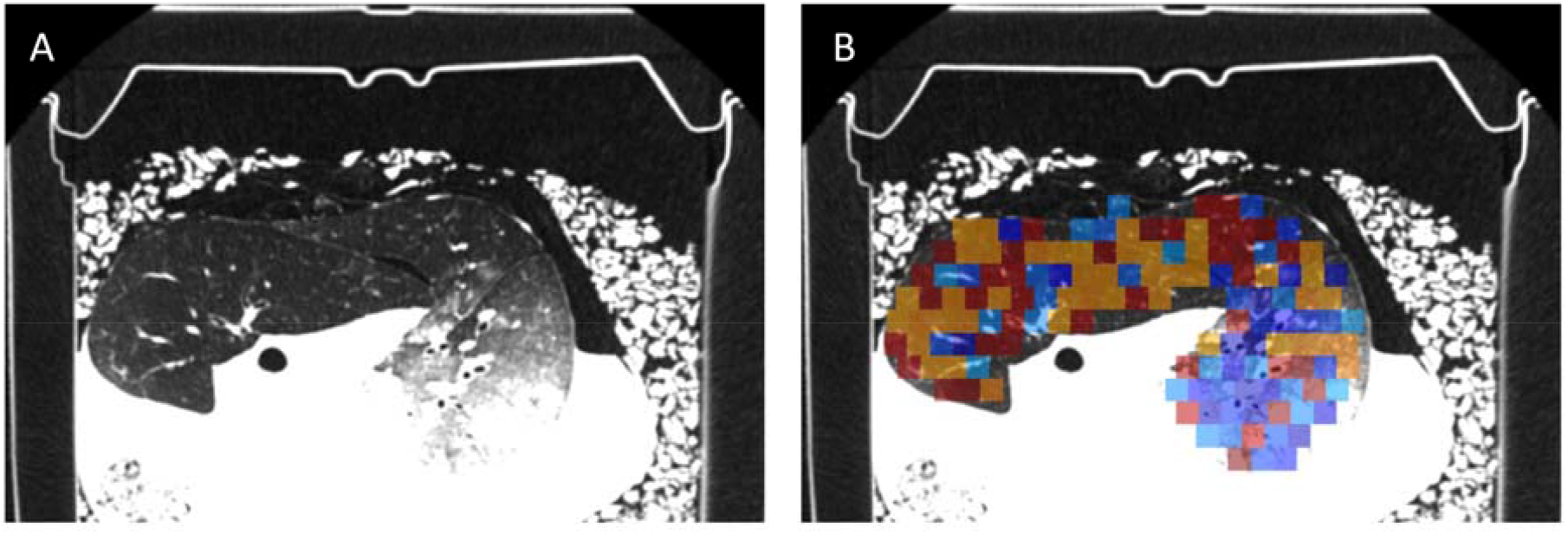
Representative CT scan with corresponding ML patch probability map from a declined donor lung identified by ML as acceptable for transplantation (False Negative). These images are from Case 10 in **Table 3**.

## Conclusions

In conclusion, this study shows the feasibility and potential to support clinicians and improve patient outcomes using this combined CT and ML strategy for donor lung screening. Results from our single center trial found that our technique was able to identify extended ICU stay and increased risk of PGD score of 3 in lung transplant recipients. In addition, we also identified donor lungs that were clinically declined but could in fact—based on our calculations—be used for transplantation, indicating an effective way to expand the donor pool.

## Data Availability

To preserve privacy and confidentiality, donor data are not available. All other data produced by the study are available upon reasonable request to the authors.

## Abbreviations

CLAD: Chronic lung allograft dysfunction
COPD: Chronic obstructive pulmonary disease
CT: Computed tomography
CVA: Cerebrovascular accident
DCD: Donation after circulatory death
FEV1: Forced expiratory volume in 1 second
FiO2: Fraction of inspired oxygen
FVC: Forced vital capacity
ICU: Intensive care unit
IPF: Idiopathic pulmonary fibrosis
ISHLT: International Society for Heart and Lung Transplantation
ML: Machine learning
PGD: Primary graft dysfunction
pO2: Partial pressure of oxygen

## Author contributions

SEV, APN and CJG conceived and designed the analysis. SR and MK developed the machine learning algorithm. SO, AV, AD, RV, LJC, AEF, DER, JV, VNL, BMV, and GMV contributed clinical or CT donor and recipient data. SR, AJB, RP, SG, SEV, and CJG assisted with analysis and interpretation. SR, SEV and CJG wrote the paper. All authors discussed and contributed to the final manuscript.

## Acknowledgements

A huge thank you to Lee Olsen, who has helped with all editing and preparation of the manuscript. Part of this work was supported by NHLBI grants R01-HL139690 and R01-HL150023 and Cystic Fibrosis Foundation grant Lama21AB0.

