## Supplemental Materials for "CT-based Machine Learning for Donor Lung Screening Prior to Transplantation"

**Supplemental Information Guide**

| **Item** | **Title/Description** |
| --- | --- |
| Supplemental Figure 1 | Workflow of Dictionary Learning Model |
| Supplemental Methods | Dictionary Learning Algorithm |
| Supplemental Results | Training and Testing ML versus SVM |
| Supplemental Figure 3 | Confusion Matrix from Test Set |
| Supplemental Table 1 | Post-Operative Pulmonary Function Measurements |

### Supplemental Figure 1

Workflow of Dictionary Learning Model

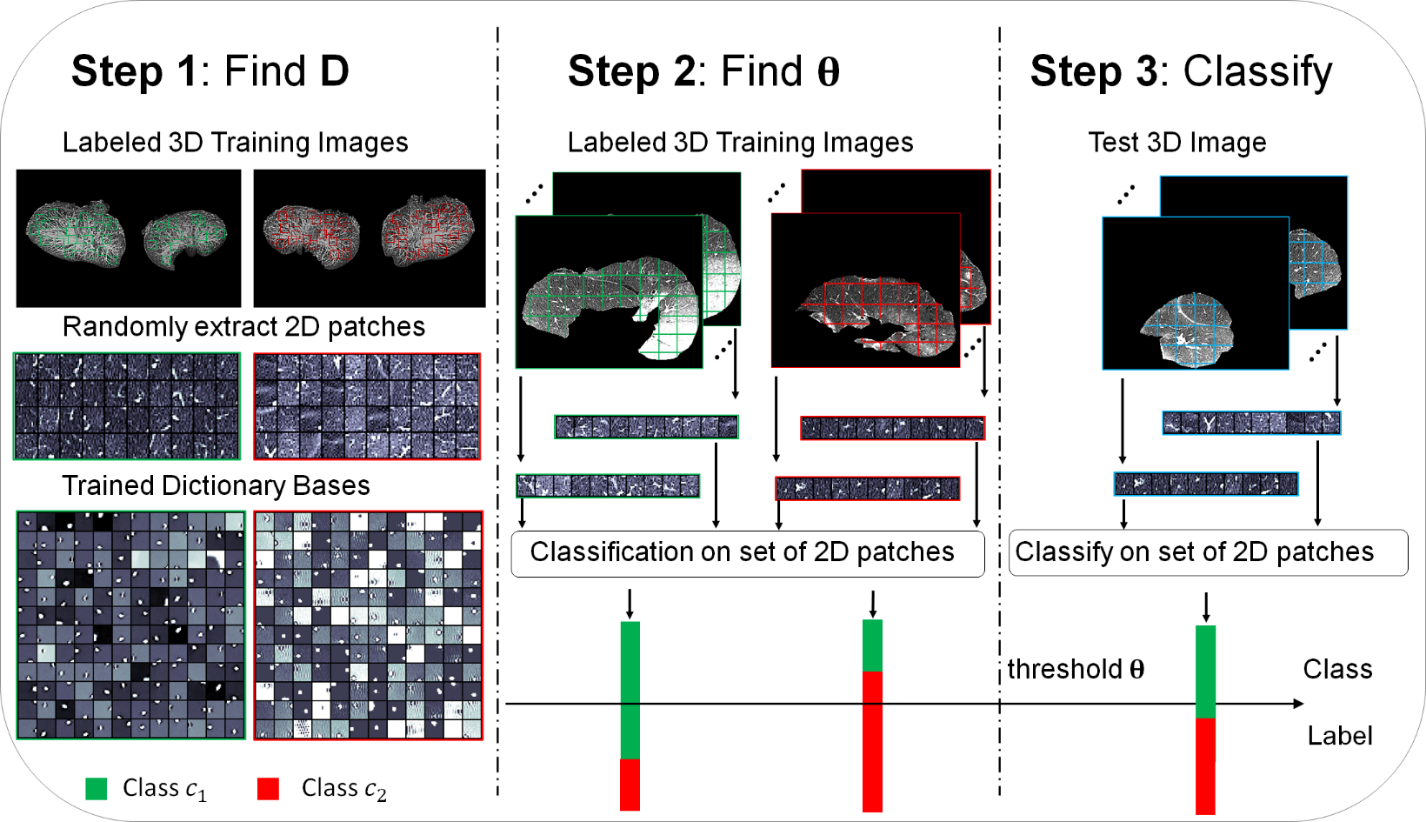

Supplemental Figure 1: Flowchart describing the various steps involved in the proposed dictionary learning algorithm. Step 1: 2D image patches are extracted from the labeled training 3D images and a class specific dictionary is trained for each class. Step 2: all 2D patches from each 3D image are classified and a threshold value is selected to classify the entire 3D image as belonging to one of the classes. Step 3: The learned class specific dictionaries and the threshold for the entire image are used to classify the test images.

### Supplemental Methods

#### Dictionary Learning Algorithm

Image sparsity has emerged as a significant property of images and sparsity-based regularization has been used for various image processing applications. ^1-8^ Sparse image representations are at the heart of many modern approaches to medical image classification and include ^9-11^. The sparse model assumes that each patch within an image can be accurately represented using a few elements of a basis set called a dictionary. For image classification problems a separate class-specific dictionary is learnt from patches belonging to each class of images. In this work, we have developed a multiview task-driven dictionary learning algorithm – a novel approach that aims to learn discriminative dictionaries for each class from multiple views of the data in a joint fashion by imposing group sparsity constraints.

*Dictionary Learning*: Our proposed method utilizes an overcomplete dictionary $\mathcal{D}$ constructed from the CT images, which is an $n \times K$ matrix whose columns represent $K$ “atoms” of size $n,$ where an “atom” is a sparse coefficient vector (i.e., a vector of weights/coefficients in the sparse basis). We train a separate dictionary for each class. Each dictionary $\mathcal{D}_{i}$ represents the image patches from class $i$ reasonably well but at the same time represents the image patches from the other classes quite poorly. There are several ways to train/learn a dictionary.^12^ In this work we have adopted the task-driven dictionary learning algorithm proposed by Mairal et al. as the basis to train our dictionaries.^13^ To train them, we solve a combinatorial optimization problem, where an approximate solution is obtained by alternating between a greedy sparse coding step using the current dictionary estimate, and a dictionary update step.

*Sparse Coding:* We assume that any image patch $x$ in a CT image can be represented as a sparse linear combination of the atoms of the dictionary $\mathcal{D}$ as: $x \approx\mathcal{D}\alpha,$ where $\alpha$ is the sparse coefficient vector. Given a dictionary, $\mathcal{D,}$ the goal in sparse coding is to find a sparse coefficient vector $\alpha.$ This requires solving a second optimization problem, the optimal solution to which is found using a greedy approach such as an orthogonal matching pursuit algorithm.^14^

*Classification:* In sparse representation-based classification, an image patch $x$ is classified according to how well the patch is represented by the class-specific dictionaries. Once a dictionary $\mathcal{D}_{i}$ has been trained for each class $i$, classification of a new image patch $x_{\mathrm{new}}$ is performed by evaluating the reconstruction/representation errors for different classes. From the class representation errors a pseudo-probability measure $P_{i}$ is computed and the image patch is assigned to the class that has the maximum probability value.

Training: The dictionary learning model was trained on a desktop workstation running a 64-bit Windows operating system (Windows 10) with an Intel Xeon W-2123 CPU at 3.6GHz with 128GB DDR4 RAM. The x-, y-, and z-dimensions of each image in our dataset was x = 512, y = 512, and z ~ 1250. Lungs from 100 donors were imaged *ex situ* using CT. A representative 2D slice of a donor lung CT image is shown in **Figure 1B** of the manuscript. The lungs within these CT images were then automatically segmented using in-house software developed using MATLAB R2020a (MathWorks, Natick, MA). Twenty donor lungs were excluded from further analysis due to poor segmentation. Our dataset for this study consists of a total 80 donor lung CT images belonging to two categories: i) 59 donor lungs that were accepted by an expert pulmonary physician and underwent lung transplantation, referred to as class 1, and ii) 21 donor lungs that were rejected and not used, referred to as class 2. We used a randomly selected subset of 14 images from both classes (7 from class 1 that were accepted for transplantation, and 7 from class 2 that were rejected) for training the dictionary learning model. The remaining 66 donor lung CT images (52 belonging to class 1 and 14 belonging to class 2) were used for testing our algorithm. A total of 2,000,000 2D image patches from the three (axial, coronal, and sagittal) views were extracted from the training data for each class to train the dictionaries. The proposed dictionary learning algorithm was developed using MATLAB R2020a (MathWorks, Natick, MA) software. We used the sparse modeling software (SPAMS) toolbox^15^ for the orthogonal matching pursuit optimization algorithm to efficiently optimize the dictionary elements. The hyper parameters of the dictionary learning algorithm include the image patch size $l,$ the number of dictionary bases $K$ for each dictionary, the sparsity controlling parameter $\lambda,$ and the positive regularization parameter $\rho$ in sparse coding. The optimal values for these parameters were automatically selected on a validation set (randomly chosen from within the training data), using the receiver operating characteristic (ROC) curves by varying one parameter at a time while keeping the others fixed and choosing that value of the parameter that maximizes the area under the curve (AUC) of the ROC curve. The parameters of the dictionary learning algorithm were set to $l=15,$ $K=250,$ $\lambda=0.001,$ and $\rho=0.001.$

### Supplemental Results

#### Training and Testing ML versus SVM

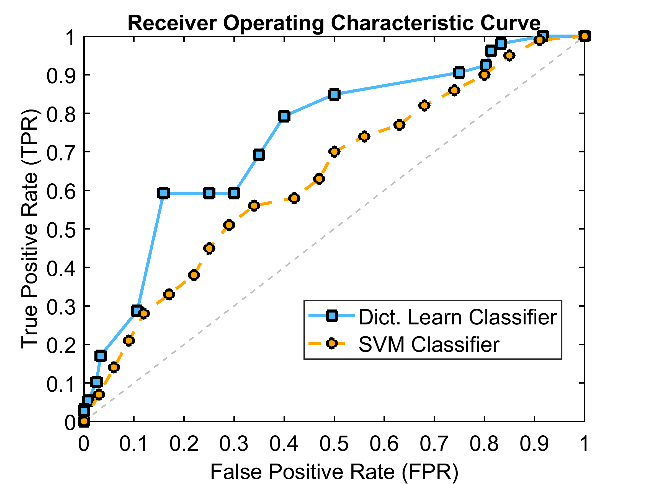

Supplemental Figure 2: Receiver operating characteristic curve for our ML model and SVM classifier.

Our algorithm was found to outperform a support vector machine (SVM) classifier when trained off 7 accepted and 7 declined donor lung CT scans. It achieved an accuracy of 72.7% with an area under the curve (AUC) of 0.743 (**Supplemental Figure 2**). In comparison, a SVM classifier was only able to achieve an accuracy of 61.8% with AUC of 0.650. Precision and recall, both measures of algorithmic performance, were also found to be higher for our algorithm over what was achieved using the SVM classifier (**Supplemental Table 2**). The optimal feature threshold using our method for classifying a complete donor lung as ‘ML Accepted’ or ‘ML Declined’ was determined using the ROC plot and found to be 0.271.

Supplemental Table 2:

| **Method** | **Precision (P)** | **Recall (R)** | **F-score** | **Accuracy [%]** | **AUC** |
| --- | --- | --- | --- | --- | --- |
| **Dictionary Learning** | 0.7801 | 0.7398 | 0.7541 | 72.74 | 0.7425 |
| **SVM Classifier** | 0.6281 | 0.7024 | 0.6632 | 61.81 | 0.6497 |

### Supplemental Figure 3

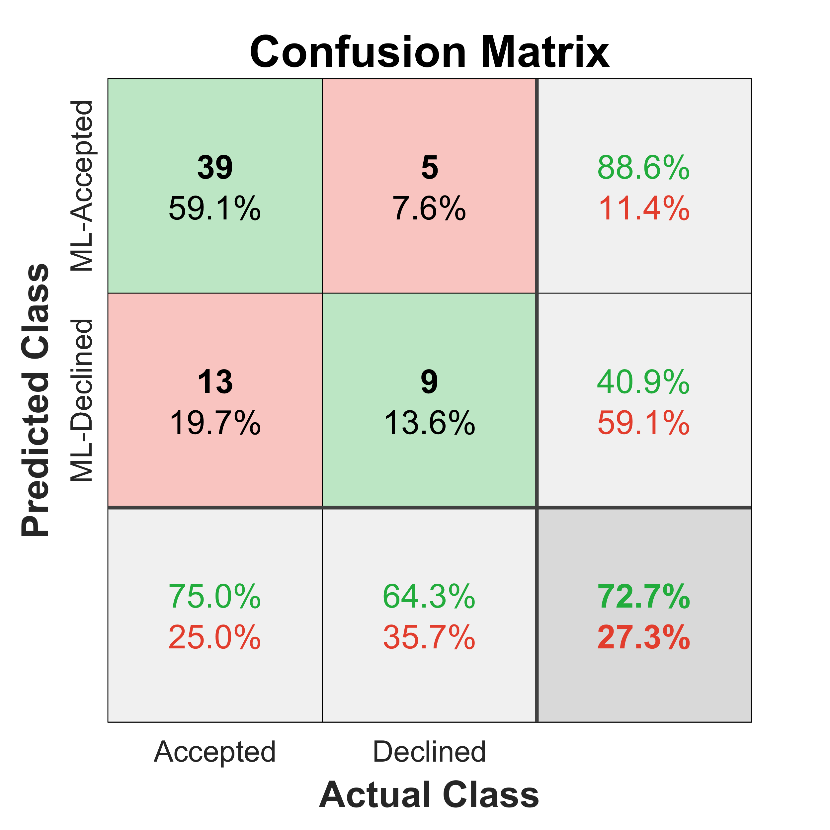

#### Confusion Matrix from Test Set

**Figure 3**: Confusion matrix provides sensitivity and specificity of ML classifications for both accepted (N=52) and declined (N=14) donor lungs from the test set. Green and Red fields in the table represent agreement and disagreement, respectively, of the ML model with clinical decision.

### Supplemental Table 1

Post-Operative Pulmonary Function Measurements

| **Recipient** | **False Negative** | **True Negative** | **P value** |
| --- | --- | --- | --- |
| Number | 13 | 38 |  |
| FEV1 Post-Operative | 2.56 (0.73) | 2.86 (0.69) | 0.140 |
| Day 30 |  |  |  |
| FEV1 (L) | 2.14 (0.67) | 2.2 (0.6) | 0.598 |
| FVC (L) | 2.52 (0.64) | 2.49 (0.73) | 0.951 |
| Day 90 |  |  |  |
| FEV1 (L) | 2.27 (0.78) | 2.49 (0.68) | 0.258 |
| FVC (L) | 2.76 (0.86) | 3.01 (0.8) | 0.317 |

Note: Data presented as mean (standard deviation). L represents liters. Group differences were determined using a Mann-Whitney U test with a significance level of p<0.05.
